# Polygenic scores clarify the relationship between mental health and gender diversity

**DOI:** 10.1101/2021.11.22.21266696

**Authors:** Taylor R. Thomas, Ashton J. Tener, Amy M. Pearlman, Katherine L. Imborek, Ji Seung Yang, John F. Strang, Jacob J. Michaelson

## Abstract

**BACKGROUND:** Gender diverse individuals are at increased risk for mental health problems, but it is unclear whether this is due to shared environmental or genetic factors.

**METHODS:** In two SPARK samples, we tested for 16 polygenic scores (PGS) effects on quantitative measures of gender diversity and mental health. In Study 1, *N* = 639 independent adults (59% autistic) reported their mental health with the Adult Self Report and their gender diversity with the Gender Self Report (GSR). The GSR has two dimensions: Binary (degree of identification with the gender opposite that implied by sex designated at birth) and Nonbinary (degree of identification with a gender that is neither male nor female). In Study 2 (*N* = 5, 165), we used categorical gender identity.

**RESULTS:** In Study 1, neuropsychiatric PGS were positively associated with mental health problems. Externalizing was positively associated with ADHD PGS (*ρ* = 0.12, *p <* 0.001, FDR = 0.10), and Internalizing was positively associated with PGS for depression (*ρ* = 0.08, *p* = 0.04, FDR = 1) and neuroticism (*ρ* = 0.11, *p* = 0.01, FDR = 0.41). Interestingly, we found no associations between gender diversity and neuropsychiatric PGS (80% powered to detect *ρ > ±*0.11). However, the GSR was positively associated with cognitive performance PGS (Binary *ρ* = 0.11, *p <* 0.001, FDR = 0.23 and Nonbinary *ρ* = 0.12, *p <* 0.001, FDR = 0.13). Binary was also positively associated with PGS for non-heterosexual sexual behavior (*ρ* = 0.09, *p* = 0.03, FDR = 0.69). In Study 2, the cognitive performance PGS effect replicated; transgender and non-binary individuals had higher PGS: *t* = 4.16, *p <* 0.001, FDR *<* 0.001. They also had higher risky behavior and anorexia PGS.

**CONCLUSIONS:** We show that while gender diversity as a trait is positively associated with mental health problems, the strongest PGS associations with gender diversity were with cognitive performance, not neuropsychiatric conditions.

## 1 INTRODUCTION

Sex and gender can have major impacts on health [1] (see Table 1 for our definitions). This stems from both extrinsic factors (e.g., healthcare barriers [2, 3]) and biological factors, with sex and gender modulating the underlying molecular mechanisms of disease and well-being [4]. In health research, sex is a more objective and well-defined variable than gender. This is because gender is often experienced on a continuum [5] and is multidimensional with binary and nonbinary dimensions. Gender can be reported through self-endorsement of categorical gender identity labels, like transgender, cisgender, nonbinary, and genderqueer. However, categorical gender identity labels may not be ideal for health research. Gender identity labels are contextually and culturally dependent (i.e., not accessible by all), and they are often nonspecific in their meanings [6]. Furthermore, gender diversity, a fundamental aspect of human diversity, is not only expressed by individuals with gender-diverse identities. People who identify as cisgender also exhibit some variation in dimensional gender diversity [7], but this diversity would be lost in studies that only use categorical gender identity labels. Therefore, parsing datasets based on numerous and nonspecific gender identity labels would erode statistical power for research studies. A continuous, multidimensional characterization of gender that uses simple and widely accessible language will enable health researchers to appropriately incorporate gender diversity.

**Table 1.**
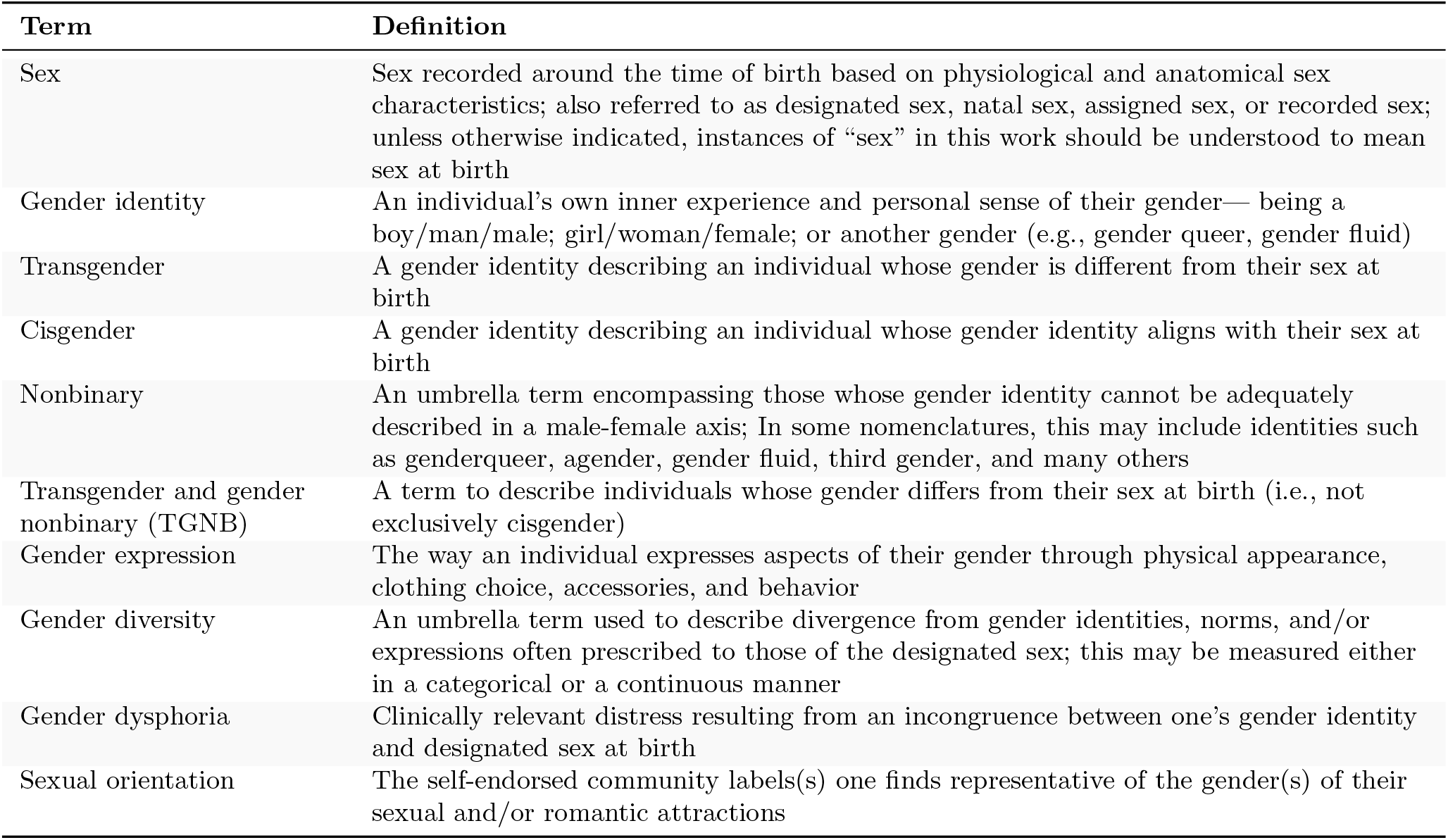
Working definitions for the gender-related terms used in this study. Some definitions are from [7] and [54].

| Term | Definition |
| --- | --- |
| Sex | Sex recorded around the time of birth based on physiological and anatomical sex characteristics; also referred to as designated sex, natal sex, assigned sex, or recorded sex; unless otherwise indicated, instances of “sex” in this work should be understood to mean sex at birth |
| Gender identity | An individual’s own inner experience and personal sense of their gender— being a boy/man/male; girl/woman/female; or another gender (e.g., gender queer, gender fluid) |
| Transgender | A gender identity describing an individual whose gender is different from their sex at birth |
| Cisgender | A gender identity describing an individual whose gender identity aligns with their sex at birth |
| Nonbinary | An umbrella term encompassing those whose gender identity cannot be adequately described in a male-female axis; In some nomenclatures, this may include identities such as genderqueer, agender, gender fluid, third gender, and many others |
| Transgender and gender nonbinary (TGNB) | A term to describe individuals whose gender differs from their sex at birth (i.e., not exclusively cisgender) |
| Gender expression | The way an individual expresses aspects of their gender through physical appearance, clothing choice, accessories, and behavior |
| Gender diversity | An umbrella term used to describe divergence from gender identities, norms, and/or expressions often prescribed to those of the designated sex; this may be measured either in a categorical or a continuous manner |
| Gender dysphoria | Clinically relevant distress resulting from an incongruence between one’s gender identity and designated sex at birth |
| Sexual orientation | The self-endorsed community labels(s) one finds representative of the gender(s) of their sexual and/or romantic attractions |

Gender diversity is a crucial variable to include in mental health research. Previous studies have reported higher rates of mental health problems in groups that express more gender diversity than the cisgender proportional majority, such as LGBTQ+ individuals [8, 9]. One study found LGBQ+ individuals had higher rates of anxiety, depression, and attempted suicide [10]. Recent work leveraging the All of Us cohort (*N* = 329, 038) found that the LGBQ+ participants had a higher prevalence of neuropsychiatric diagnoses [11]. The exact mechanisms are not fully understood. However, research has shown that poorer mental health is at least partially due to factors related to the experienced adversity from sexual orientation and/or gender diversity. One study found that discrimination and resilience partially mediated negative mental health outcomes in LGBTQ+ college students [12]. Another study found that access to gender-affirming hormone therapy for transgender and gender nonbinary youth was associated with reduced risk of depression and suicidality [13]. To our knowledge, no study has used genetic data, so any possible genetic mechanisms are unknown.

Most behaviors are somewhat heritable, so we and others have hypothesized that gender identity and gender diversity are also susceptible to genetic influences. [14]. One twin study of *N* = 4, 426 females estimated the heritability of adult gender expression (i.e., self-reported masculinity and femininity) at 11%, and retrospective childhood gender typicality at 31% [15]. However, the searches for specific loci have been underpowered for gene discovery [16, 17]. Genome-wide association studies (GWAS) of human behavior often uncover many associated loci that each have a small effect size and contribute additively [18]. Loci associated with one trait are often associated with other traits, which suggests that the two traits have a degree of pleiotropy. However, the predictive power of the PGS depends on the GWAS power, which is driven chiefly by sample size. Among the well-powered GWAS, the most proximal trait to gender diversity is the non-heterosexual sexual behavior (NHSB) GWAS [19] performed in *N* = 408, 995 UK Biobank participants. The trait was defined as yes/no response to ever having sex with someone of the same sex (the nuance between same-sex versus same-gender is lost due to the nature of the question). The estimated heritability of NHSB ranged from 8% to 25%. It was positively genetically correlated with several neuropsychiatric conditions and personality traits. However, the interpretation of these genetic correlations is limited because of the confounding with experienced adversity and psychiatric diagnoses.

In this study, we investigated whether gender diversity, like NHSB, is genetically associated with other behaviors and if this plays a role in mental health. We invited a subset of participants from SPARK [20] to complete surveys about their mental health and gender identity. SPARK is a national genetic study totalling more than 300,000 participants with and without autism. Previous studies have shown that there is an enrichment of gender diversity in autism [21], and gender diverse individuals were found to have increased levels of clinically relevant autistic traits and increased likelihoods of autism diagnoses [22]. This makes SPARK a logical choice for investigating this topic. In our sample of *N* = 696 (*N* = 639 of European genetic ancestry), we calculated PGS for 16 behavior traits. We also administered two psychometrically valid self-report tools. The first, the Adult Self Report (ASR) [23], measures several mental health outcomes and adaptive behaviors. The second, the Gender Self-Report [24] captures two quantitative dimensions of gender diversity: Binary Gender Diversity, the extent one experiences themselves as the other binary gender (i.e., different from their sex designated at birth), and Nonbinary Gender Diversity, the extent one experiences themselves as neither female nor male. We then sought to answer the following questions: First, are the ASR scores phenotypically associated with the GSR scores? Second, are behavior-related PGS associated with ASR and GSR scores? How are the PGS associations different for the ASR versus the GSR? Lastly, do the PGS findings broadly replicate in a larger sample with a categorical gender identity phenotype instead of the GSR? See Figure 1 for an overview of the study.

**Figure 1.**
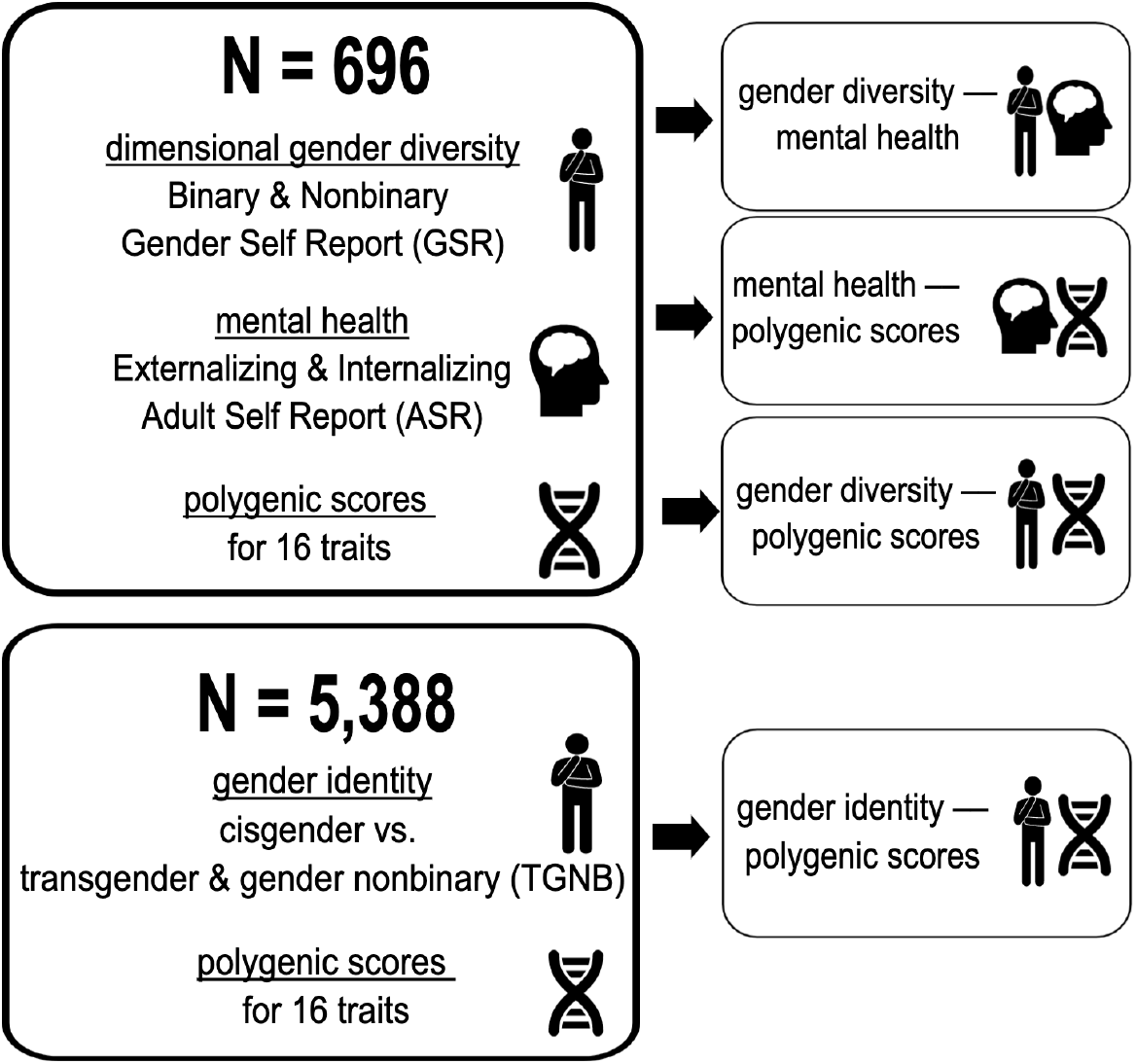
Overview of the study. In *N* = 696 (*N* = 639 of European genetic ancestry), dimensional gender diversity was measured using the Gender Self Report (GSR) and mental health was measured using the Adult Self Report (ASR). The GSR and ASR scores were then tested for associations with 16 polygenic scores (PGS) for psychiatric diagnoses, personality, and cognition. We then used categorical gender identity in *N* = 5, 388 (*N* = 5, 165 of European genetic ancestry) to test for the replication of these PGS associations in the larger sample.

## 2 RESULTS

### 2.1 Phenotypic associations between gender diversity and mental health

The demographic characteristics of the SPARK Research Match participants are shown in Table 2. The final sample size was *N* = 696 with *N* = 639 of European genetic ancestry. Approximately one third of the cohort identified as transgender or gender nonbinary (TGNB). Fifty-eight percent of the participants were autistic and 22% were male.

**Table 2.**
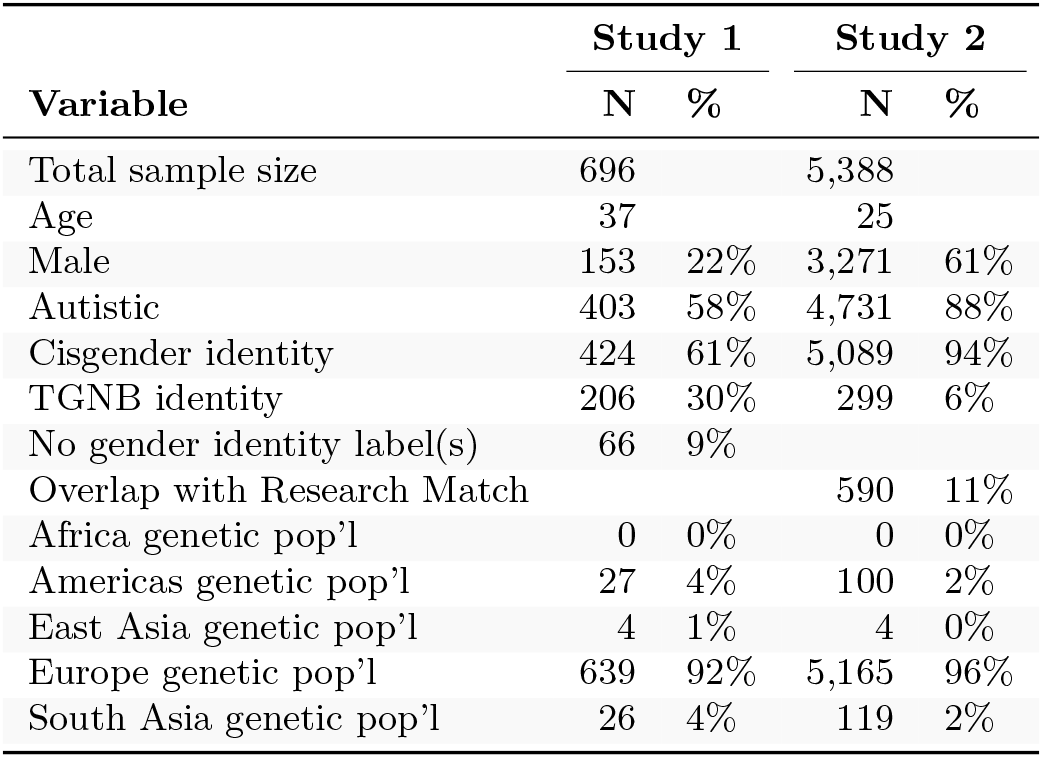
Participant Demographics. Study 1 used the two continuous measures of gender diversity from the Gender Self Report, and Study 2 used categorical gender identity.

The two gender diversity scores, Binary and Nonbinary Gender Diversity, were from the Gender Self Report (GSR) factor analysis [24] . GSR scores range from 0 (no gender diversity) to 1 (high gender diversity), with the modes near 0(Figure S1. The scores were adjusted for age, sex designated at birth, and autism by linear regression residualization, and then Z-scaled (*µ* = 0, *σ* = 1). The distributions of these two scores are shown in Figure 2A and are colored by sexual orientation and gender identity. The general trend shows higher gender diversity in LGBQ+ and TGNB participants. Binary and Nonbinary were positively correlated: *ρ* = 0.57, *p <* 0.001 (Figure 2B).

**Figure 2.**
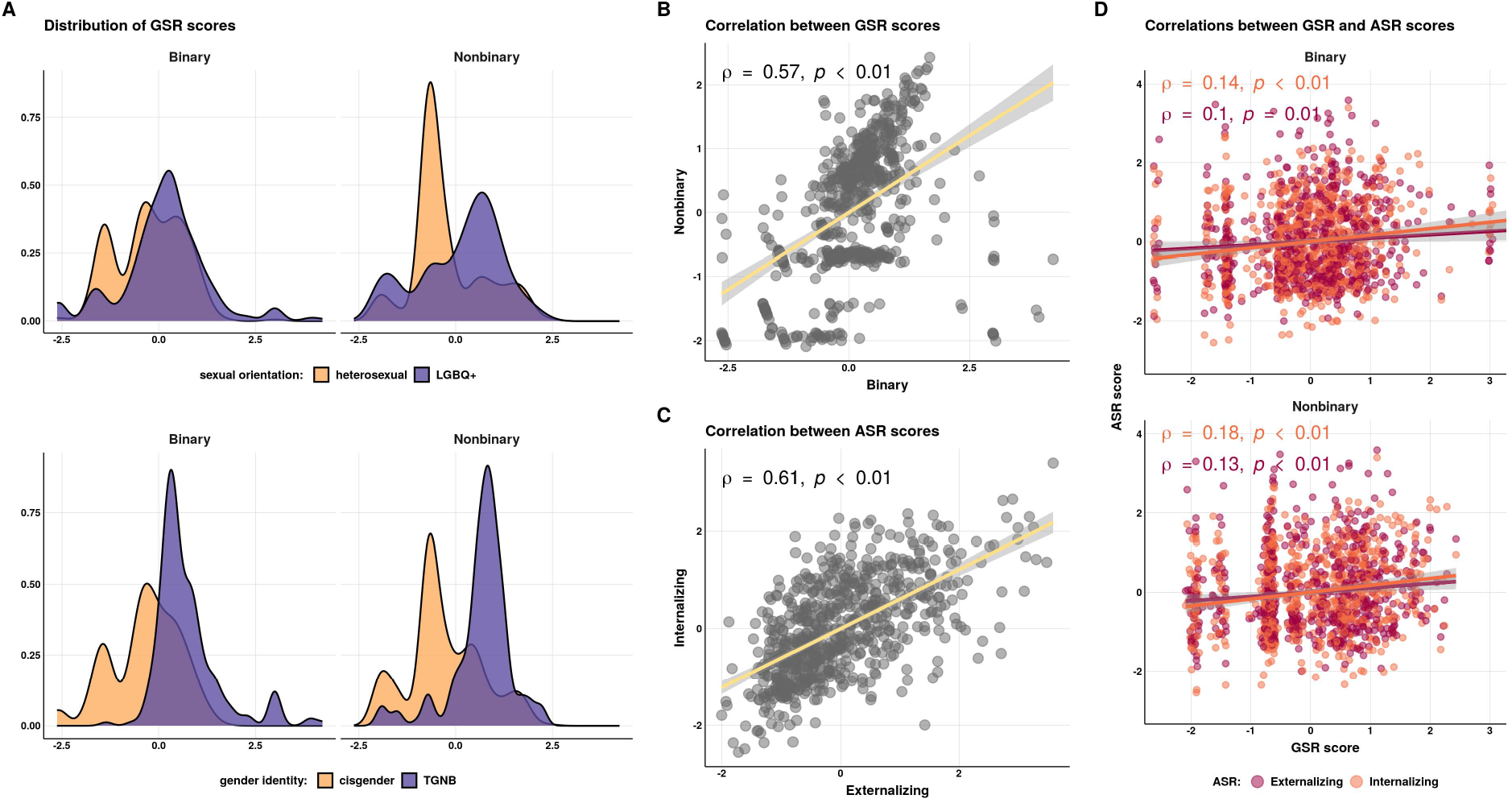
Distributions and correlations of the Gender Self Report and Adult Self Report scores. **(A)** Distribution of the two dimensional gender diversity measures from the Gender Self Report (GSR) GSR: Binary and Nonbinary Gender Diversity. The sample size was *N* = 696. The GSR scores were adjusted for age, sex, and autism. The histograms are colored by self-endorsed sexual orientation labels (top panel) and gender identity labels (bottom panel). Distributions of the GSR scores before adjusting for age, sex, and autism are shown in Figure S1. **(B)** Correlation of the two GSR scores. **(C)** Correlation of the two mental heath measures from the Adult Self Report (ASR): Externalizing and Internalizing problems. The ASR scores were also adjusted for age, sex, and autism. **(D)** Correlations between the GSR scores and ASR scores.

The two mental health scores, Externalizing and Internalizing, were from the Adult Self Report (ASR) [23]. The scores were adjusted for age, sex designated at birth, and autism by linear regression residualization and then Z-scaled. Externalizing and Internalizing were positively correlated: *ρ* = 0.61, *p <* 0.001 (Figure 2C). The ASR scores were positively correlated with the GSR scores (Figure 2D). Binary was more correlated with Internalizing (*ρ* = 0.14, *p <* 0.001) than Externalizing (*ρ* = 0.10, *p* = 0.01). Nonbinary was also more correlated with Internalizing (*ρ* = 0.18, *p <* 0.001) than Externalizing (*ρ* = 0.13, *p <* 0.001).

### 2.2 Polygenic score associations with gender diversity and mental health

We next assessed the associations between GSR and ASR scores with 16 polygenic scores (PGS) of behavior (Figure 3). The PGS were adjusted for the 20 principal genetic components and then age, sex designated at birth, and autism. Correlations were run in the European subset (*N* = 639). The sample size of *N* = 639 has 80% power at *α* = 0.05 to detect effects *ρ > ±*0.11, meaning that the absence of significant effects must be carefully interpreted. In addition to these partial correlations, we ran linear regression with age, sex, and autism included as covariates (Table S1). Multiple testing corrections for the 16 PGS were performed with the Benjamini and Yekutieli method [25], which is a conservative method for multiple testing correction.

**Figure 3.**
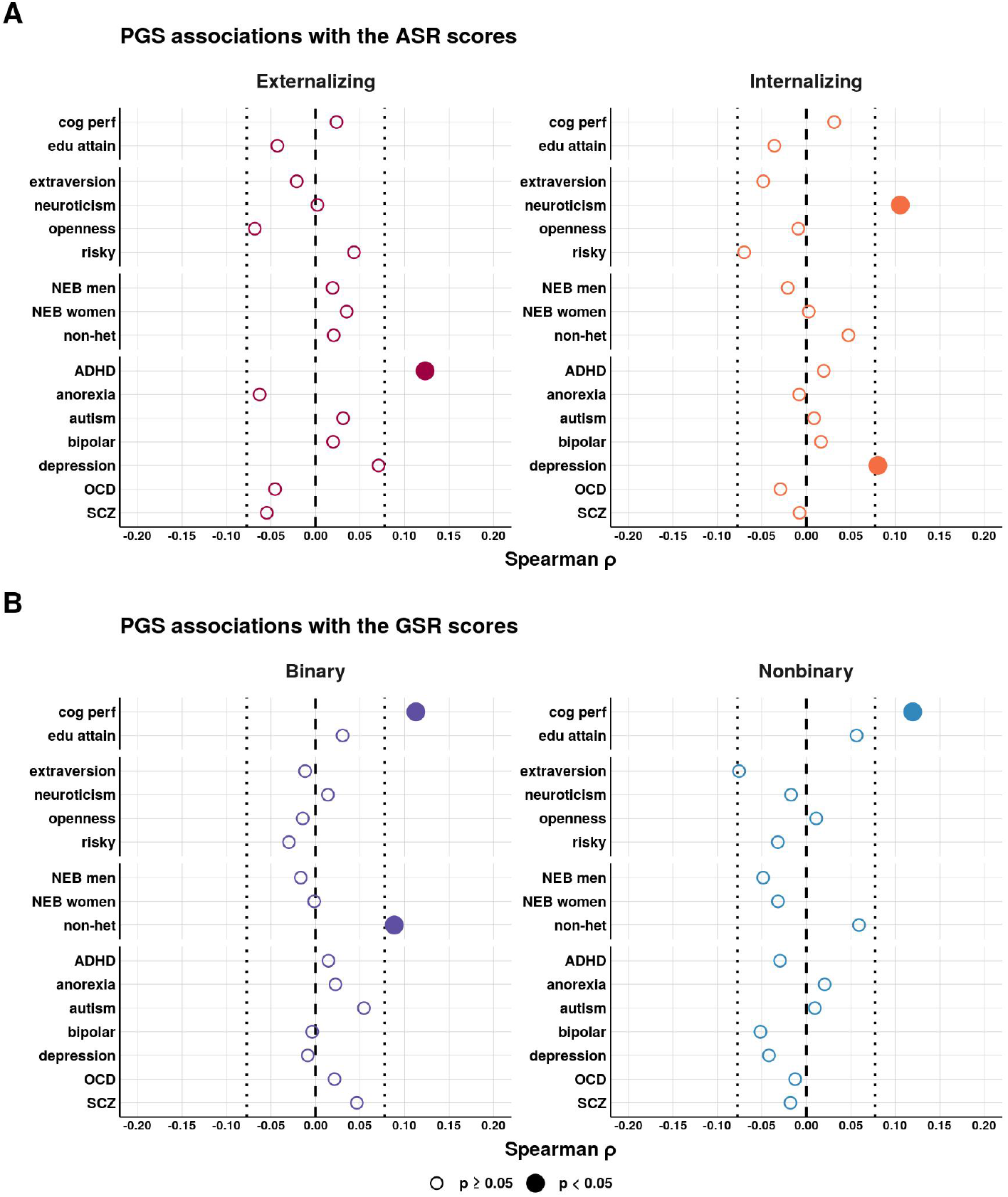
Polygenic score associations with the Gender Self Report and Adult Self Report scores. Polygenic score (PGS) associations with the **(A)** Adult Self Report (ASR) scores and **(B)** Gender Self Report (GSR) scores. The PGS, as well as the GSR and ASR phenotypes, were adjusted for age, sex, and autism prior to correlations by linear regression residualization. The correlations shown here were run in the European subset (*N* = 639). Correlations were also run stratified by autism (Figure S3). The PGS linear model associations with age, sex, and autism included as covariates (as opposed to residualizing the covariates from the variables) are shown in Table S1. The partial correlations not filtered on genetic ancestry (*N* = 696) are shown in Figure S2. Multiple testing corrections for the 16 PGS were performed with the Benjamini and Yekutieli method [25]. PGS abbreviations: **risky** = risky behavior, **NEB** = number of children ever born, **non-het** = non-heterosexual sexual behavior, **SCZ** = schizophrenia.

The ASR scores had unsurprising correlations with psychiatric PGS. Externalizing was positively correlated with ADHD PGS (*ρ* = 0.12, *p <* 0.001, *FDR* = 0.10). Internalizing was positively correlated with depression PGS (*ρ* = 0.08, *p* = 0.04, *FDR* = 1) and neuroticism PGS (*ρ* = 0.11, *p* = 0.01, *FDR* = 0.41).

As expected, non-heterosexual sexual behavior (NHSB) PGS was positively correlated with Binary: *ρ* = 0.09, *p* = 0.03, *FDR* = 0.69. NHSB PGS was also positively correlated with Nonbinary, although the correlation did not reach nominal significance: *ρ* = 0.06, *p* = 0.14, *FDR* = 1. Strikingly, cognitive performance PGS was significantly positively correlated with Binary (*ρ* = 0.11, *p <* 0.001, *FDR* = 0.23) and Nonbinary (*ρ* = 0.12, *p <* 0.001, *FDR* = 0.13), meaning that the polygenic propensity for higher cognitive performance was associated with elevated binary and nonbinary gender diversity. No psychiatric PGS were significantly correlated with the GSR scores, although autism PGS approached nominal significance with Binary (*ρ* = 0.05, *p* = 0.17, *FDR* = 1).

The results were comparable when not filtering on genetic ancestry (*N* = 696, see Figure S2). Cognitive performance PGS was positively correlated with Binary *ρ* = 0.09, *p* = 0.02, *FDR* = 0.66) and Nonbinary (*ρ* = 0.11, *p <* 0.001, *FDR* = 0.16). NHSB PGS was positively correlated with Binary: *ρ* = 0.09, *p* = 0.02, *FDR* = 0.66. Autism PGS was positively correlated with Binary: *ρ* = 0.08, *p* = 0.05, *FDR* = 0.82. We tested whether the correlations trended in the same direction when run separately in the autistic subset (*N* = 376), versus those without an autism diagnosis (*N* = 263) (Figure S3). These tests were not well powered, but cognitive performance PGS was positively correlated with Binary in the autistic subset (*ρ* = 0.10, *p* = 0.04, *FDR* = 1) and the non-autistic subset (*ρ* = 0.13, *p* = 0.04, *FDR* = 1), as well as Nonbinary in the autistic subset (*ρ* = 0.12, *p* = 0.02, *FDR* = 1) and the non-autistic subset (*ρ* = 0.08, *p* = 0.19, *FDR* = 1).

### 2.3 Replication of polygenic score associations with categorical gender identity

We next tested if the PGS associations with the GSR were similar in a larger sample (*N* = 5, 388, *N* = 5, 165 of European genetic ancestry) with a categorical gender identity phenotype (Study 2). We used the background history files provided by SPARK to label individuals as cisgender or TGNB by using discordance between the participant’s designated sex at birth (options: Male or Female) and their gender (options: Male, Female, or Other) to categorize the participants as TGNB or cisgender. We merged this with Study 1 for a final sample size of *N* = 5, 388, with *N* = 590 from Study 1 and the remaining *N* = 4, 798 from the background history reporting. The mean age for Study 2 was 25 years and 88% of the participants were autistic (Table 2).

We tested for PGS differences between the two gender identity groups with t-tests (Figure 4). The strongest effect was observed for cognitive performance PGS, with the TGNB group being significantly higher (*x_d_* = 0.26 [0.14, 0.39], *t* = 4.16, *p <* 0.001, *FDR <* 0.001). The TGNB group also had significantly higher PGS for risky behavior (*x_d_* = 0.12 [0.01, 0.23], *t* = 2.12, *p* = 0.03, *FDR* = 0.67) and anorexia (*x_d_* = 0.12 [0.01, 0.24], *t* = 2.09, *p* = 0.04, *FDR* = 0.67). NHSB PGS was close to nominal significance (*x_d_* = 0.11 [*−*0.01, 0.23], *t* = 1.86, *p* = 0.06, *FDR* = 0.84).

**Figure 4.**
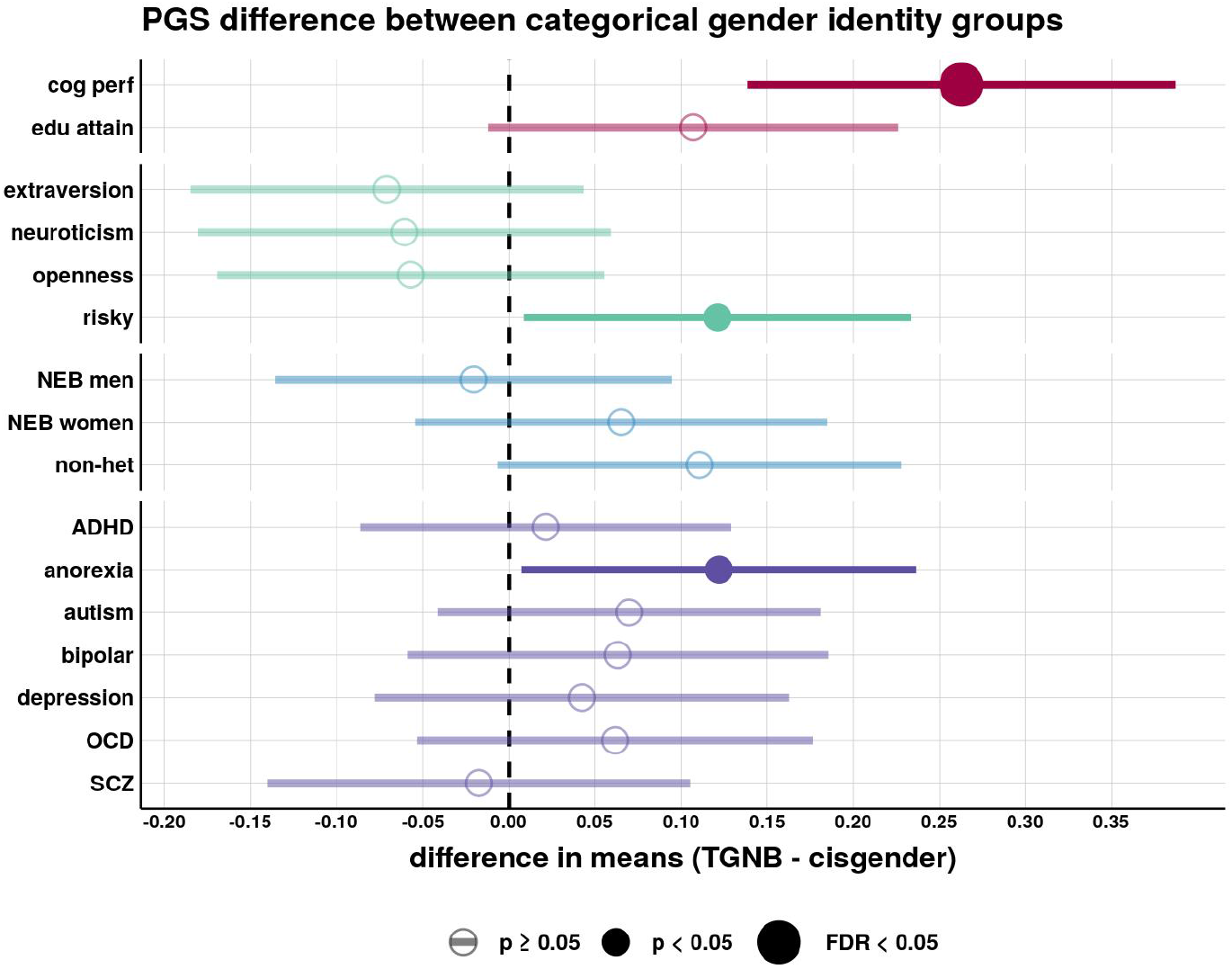
Replication of polygenic score associations with categorical gender identity in a larger sample (Study 2). Polygenic score (PGS) difference in means (t-tests) between gender identity groups in the larger cohort of European genetic ancestry (*N* = 5, 165). The two gender identity groups are cisgender (*N* = 4, 879) versus transgender and gender nonbinary (TGNB, *N* = 286). The PGS were adjusted for age, sex, and autism prior to performing the t-tests. The results with the PGS not adjusted for age, sex, and autism are shown in Figure S5. The results not filtered on genetic ancestry (*N* = 5, 388) are shown in Figure S4. Multiple testing corrections for the 16 PGS were performed with the Benjamini and Yekutieli method [25]. PGS abbreviations: **risky** = risky behavior, **NEB** = number of children ever born, **non-het** = non-heterosexual sexual behavior, **SCZ** = schizophrenia.

We repeated the tests not filtering on genetic ancestry (*N* = 5, 388) (Figure S4). The strongest effect was still cognitive performance PGS, which was higher in the TGNB group (*x_d_* = 0.24 [0.12, 0.36], *t* = 3, 86, *p <* 0.001, *FDR* = 0.01). The TGNB group was also higher for risky behavior PGS (*x_d_* = 0.12 [0.01, 0.23], *t* = 2.09, *p* = 0.04, *FDR* = 0.79) and NHSB PGS (*x_d_* = 0.12 [0, 0.23], *t* = 2.02, *p* = 0.04, *FDR* = 0.79). We repeated the tests by adjusting the PGS for only age (Figure S5A) and not adjusting the PGS for age, sex, or autism (Figure S5B).

### 2.4 Interactions between gender diversity, mental health, and polygenic scores

Although our smaller sample size was not well-powered for interactions, having found little evidence for main effect PGS associations that explained the mental health associations with gender diversity, we decided to investigate interactions between PGS and gender diversity that could potentially explain mental health outcomes. We tested for PGS-by-GSR interactions with three linear models and performed correlations stratified by PGS (Figure S6). The first model included the covariates (age, sex, and autism) as main effects and interactions with PGS and GSR, as recommended by [26]. For the second model, we maintained consistency with the previous analysis that adjusted for covariates prior to tests. For the third model, we binarized the PGS into the upper quartile (coded as 1) and the lower quartile groups (coded as 0), with *N* = 160 in each group and the middle 50% removed. From the third model, we identified three nominally significant PGS-by-GSR interactions, specifically the schizophrenia and depression PGS. The most prominent interaction was Nonbinary and schizophrenia PGS interaction on Internalizing (*β* = 0.32, [0.1, 0.53], *p <* 0.001, *FDR* = 0.20). Within the entire sample of *N* = 639, Nonbinary and Internalizing were positively correlated: *ρ* = 0.18, *p <* 0.001. However, this apparent main effect appears to be driven by a context-specific interaction with PGS: in the subset at highest schizophrenia PGS (e.g., the upper quartile, *N* = 160), the correlation between Nonbinary and Internalizing was *ρ* = 0.36, *p <* 0.001. While in the lowest risk group (e.g. the lower quartile, *N* = 160), there was no correlation: *ρ* = 0.02, *p* = 0.81. The interaction between depression PGS and Nonbinary on Internalizing was also similar (*β* = 0.23, [0.02, 0.45], *p* = 0.04, *FDR* = 0.97).

## 3 DISCUSSION

Our analyses were the first to address the relationships of multidimensional gender diversity with mental health and genetics. We used two quantitative measures of gender diversity, Binary and Nonbinary Gender Diversity, from the Gender Self-Report (GSR) in a neurodiverse sample of *N* = 696 adults in SPARK [20]. In our sample, we found greater gender diversity in female, autistic, and LGBTQ+ participants. Due to the structure of SPARK recruitment, we were only able to collect data from independent adults with autism or non-autistic immediate family members of someone with autism (mainly parents). Therefore, the elevated gender diversity in the autistic participants should be interpreted with the caveat that the non-autistic participants were older and presumed to adhere to more traditional gender roles. However, these results are in line with previous research showing the enrichment of gender diversity in autism [21]. Intriguingly, while our results showed higher gender diversity in LGBTQ+ participants, many people who were cisgender also showed evidence of gender diversity, though not enough to report TGNB identities. This underscores the value of the GSR in capturing dimensional gender diversity beyond self-endorsed identities alone.

We tested 16 behavior-related PGS for association with the two GSR dimensions, and strikingly, the strongest association was cognitive performance PGS positively associated with both Binary and Nonbinary (Figure 3A). This finding was validated in our larger sample of *N* = 5, 165 with categorical gender identity; cognitive performance PGS was higher in the TGNB group compared to the cisgender group (Figure 4). This suggests that cognitive capacity may be an important component in the development of more complex and nuanced gender identities. Beyond cognitive performance, non-heterosexual sexual behavior (NHSB) PGS was positively correlated with Binary. Although gender identity and sexual orientation are distinct, the NHSB GWAS is a well-powered GWAS that is adjacent to gender diversity. Recent research has found that just within heterosexuals, the NHSB PGS is positively associated with an increased number of partners [27]). Building on this, our results suggest that gender diversity may be part of a pleiotropic ensemble of traits with adaptive advantages (e.g., cognitive performance).

We expected neuropsychiatric PGS to also be positively correlated with the GSR, considering NHSB is positively genetically correlated with several neuropsychiatric conditions [19]. Our sample size was on the low end for PGS associations: *N* = 639 provides 80% power for detecting *ρ ±* 0.11, meaning we are not powered to detect small effects. However, in light of this previous research, it was surprising that we found no strong significant positive associations of neuropsychiatric PGS with the GSR. This suggests that, within the statistical power limits of our sample, gender diversity may not have a strong direct genetic relationship with adult-onset psychiatric disorders. Instead, in our sample higher gender diversity had the strongest genetic relationships with higher cognitive ability and NHSB.

The lack of a main genetic effect that links psychiatric conditions and gender diversity, combined with our observation that the GSR scores nevertheless show numerous significant correlations with poorer self-reported mental health prompted us to examine the possibility of a relationship between gender diversity and mental health that depends on the level of genetic risk (i.e., an interaction between polygenic risk and gender diversity). We observed differences in correlations when stratifying by schizophrenia and depression PGS (Figure S6D). Groups with high depression and schizophrenia PGS had the strongest GSR-ASR correlations, whereas the GSR-ASR correlations in the low PGS groups were absent (i.e., not nominally significant). This suggests that PGS for depression and schizophrenia might interact with gender diversity (or related environmental factors such as discrimination and/or minority stress), ultimately affecting mental health. In other words, the observed relationship between gender diversity and mental health may not be solely environmental or genetic, but rather an interaction of the two. However, our sample size limits the ability to detect PGS main effects, let alone interactions, and therefore the interaction effects must be interpreted with the understanding that they are small and not significant after multiple testing correction.

Our results and their interpretations have several limitations. The primary limitation is the small sample size, and we were only powered to detect strong PGS effects. In addition, age, sex designated at birth, and autism are entangled with other variables of interest. Autism is confounded at the genetic level, as observed in previous work showing educational attainment [28] and cognitive performance [29] are positively genetically correlated with autism. However, we repeated our analyses without adjusting the PGS and phenotypes for autism (Table S1 and Figure S5) and also stratifying by autism (Figure S3) and found the results to be robust against the inclusion or omission of autism.

In summary, our findings show that gender diversity, as captured by the GSR, has dimensional properties that share common genetic factors with cognitive performance and NHSB. In agreement with previous studies, we find that higher gender diversity is correlated with poorer mental health, but our results suggest that any polygenic contribution of psychiatric risk alleles to gender diversity, if such contributions exists, are not large. Rather, one’s polygenic background may function as a risk/resilience mechanism that interacts with gender diversity (and/or the adversity that comes with it) in shaping mental health outcomes.

## 4 METHODS AND MATERIALS

### 4.1 Sample description

SPARK [20] is a nationwide autism study in the United States involving more than 300,000 participants, with genetic data available for many of the participants. SPARK is approved by the Western IRB (#20151664). For Study 1, independent adults, with or without autism, were invited to participate in our SPARK Research Match. The Research Match was approved by the University of Iowa Institutional Review Board (IRB #201611784). Those consented to participate were asked to complete the Gender Self Report (GSR) [24], the Adult Self Report (ASR) [23], and additional questions on sexual orientation, gender identity, and gender expression. The sample size was *N* = 818. We removed nine individuals who had withdrawn from SPARK since the Research Match based on Version 8 (*N* = 809). *N* = 696 was the final sample size after genetic data availability and quality control filtering. For Study 2, we used the Version 8 background history files. The independent adult data was self-report, whereas the child and siblings data were parent report. We kept children 14 years or older and whose cognitive impairment status at enrollment was not significantly below age.

### 4.2 Study 1 phenotypes

#### Labels of gender identity and sexual orientation

Participants were able to select as many labels for gender identity and sexual orientation they found applicable. Selections of nonbinary, demigender, gender fluid, third gender, agender, gender neutral, pangender, bigender, and gender queer were categorized as nonbinary/neutral. Cisgender and transgender were each categorized separately. Participants who did not endorse any of the listed gender identities were excluded from analyses using gender identity labels (*N* = 66 of *N* = 696). For sexual orientation, participants selecting lesbian, gay, bisexual, pansexual, homosexual, queer, and/or polysexual were grouped as LGBQ+ and heterosexual orientation was classified separately. Participants who did not select any of the listed sexual orientation labels were excluded from analyses using sexual orientation labels (*N* = 73 of *N* = 696).

#### Gender Self Report (GSR)

The GSR itemset was developed through an iterative multi-input community driven process with autistic cisgender, autistic gender-diverse, and non-autistic cisgender and gender-diverse collaborators [24]; Open Science Framework Development Summary: https://osf.io/qh25d/?view_only=c0ce41d07bca4af1b792e074d51b7ded. The final GSR itemset is composed of 30 questions. The GSR factor analysis and generation of Binary and Nonbinary factor scores are described in [24]. In the genetic sample of *N* = 696, the GSR scores were adjusted for age, sex designated at birth, and autism by linear regression residualization and then Z-scaled.

#### Adult Self Report (ASR)

The ASR [23] is a questionnaire of 129 items assessing a range of adaptive behaviors and mental health outcomes. From *N* = 809, five participants were removed because they had 12 (approximately 10%) or more missing ASR items. In the remaining *N* = 804, 0.2% of the data was missing, with no item having more than five missing data points. The missing data was imputed to the median. The two ASR subscales were Externalizing and Internalizing. Externalizing is a composite score of aggressive, rulebreaking, and intrusive behavior, and Internalizing is a composite score of anxious, withdrawn depressed, and somatic complaints. In the genetic sample of *N* = 696, the ASR scores were adjusted for age, sex designated at birth, and autism by linear regression residualization and then Z-scaled.

### 4.3 Study 2 phenotypes

If the participant’s designated sex at birth (options: Male or Female) did not match their gender (options: Male, Female, or Other) then the participant was classified as TGNB. We then merged with our Research Match gender identity labels. The final sample size was *N* = 5, 388 with *N* = 590 from the Research Match and the other *N* = 4, 798 from the background history.

### 4.4 Genotype quality control and imputation

We used the genotype array data from SPARK integrated whole-exome-sequencing (iWES1) 2022 Release and the SPARK whole-genome-sequencing (WGS) Release 2, 3, and 4. iWES1 (*N* = 69, 592) was quality controlled on release, including removing samples due to heterozygosity or high missingness, so no further quality control was performed by us before genotype imputation. iWES1 provided genetic ancestry assignments based on the 1000 Genomes populations [30]. WGS Release 2 (*N* = 2, 365), Release 3 (*N* = 2, 871), and Release 4 (*N* = 3, 684) were not quality controlled on release, so we performed quality control using PLINK [31] before genotype imputation. First, we removed participants from the WGS releases if they were in iWES1. Second, we removed variants with missingness higher than 0.1 and participants with missingness higher than 0.2. Third, we merged the three releases and removed any participant whose heterozygosity (F statistic) was not within 3 standard deviations of the mean heterozygosity across the three releases. We then used the TopMed reference panel [32] to identify strand flips. The final sample size for WGS 2-4 was *N* = 8, 152. iWES1 and WGS 2-4 were then imputed to the TopMed [32] reference panel using the Michigan Imputation Server [33] with the phasing and quality control steps included and to output variants with imputation quality r2 *>* 0.3. After imputation, the variants were filtered to only the HapMap SNPs (*N* = 1, 054, 330 variants) with imputation quality r2 *>* 0.8 using bcftools [34]. They were lifted over from hg38 to hg19 using the VCF-liftover tool (https://github.com/hmgu-itg/VCF-liftover) and the alleles normalized to the hg19 reference genome. Finally, the files were merged and only variants with 0% missingness were retained (*N* = 914, 328).

### 4.5 Genetic ancestry

Genetic principal components (PCs) were calculated using the bigsnpr package [35], specifically following the author’s recommendations [36] and their tutorial: https://privefl.github.io/bigsnpr/articles/ bedpca.html. In summary, we 1.) used the snp_plinkKINGQC function to identify and remove related participants at the KING threshold of 2^−3.5^, 2.) performed PC analysis using the bed_autoSVD on just the unrelated participants, 3.) detected PC outliers and removed them, 4.) recalculated the PCs, and 5.) projected the PCs onto the entire cohort using the bed_projectSelfPCA function. We used the 40 PCs and performed k-means clustering with K = 5 (for the five populations of 1000 Genomes [30]) and used the genetic ancestry labels from iWES1 to assign labels to the genetic population clusters.

### 4.6 Relatedness

For Study 1, from the *N* = 809 Research Match participants whom completed the GSR, *N* = 804 completed the ASR and *N* = 727 had genetic data. This subset was pruned to remove related participants using GCTA [37] with a relatedness threshold of 0.125, corresponding to approximately third-degree relatives (*N* = 31 removed). For Study 2, we retained only one participant from each family, with prioritization towards TGNB identities and then removed related participants at the same threshold.

### 4.7 Polygenic scores

PGS were calculated using LDpred2 [38] and the bigsnpr tools [35] in R [39]. Because SPARK is family-based, an external LD reference based on *N* = 362, 320 in the UK Biobank (provided by the authors of LDpred2) was used to calculate the genetic correlation matrix, estimate heritability, and calculate infinitesimal beta weights. PGS were calculated from the following genome-wide association studies (GWAS): ADHD [40], anorexia nervosa [41], autism [28], bipolar disorder [42], major depression [43], OCD [44], schizophrenia [45], cognitive performance [46], educational attainment [46], and non-heterosexual sexual behavior (NHSB) [19]. The public LDpred2 beta weights from the Polygenic Index Repository [47] were used to calculate PGS for extraversion [48], neuroticism [49], openness [50], risky behavior [51], number of children ever born (men) [52], and number of children ever born (women) [52]. To account for genetic ancestry, we residualized the 20 genetic PCs from the PGS. We also accounted for age, sex designated at birth, and autism using linear regression residualization. Lastly, the PGS were Z-scaled. We performed PGS processing separately for Study 1 (*N* = 696) and Study 2 (*N* = 5, 388).

PGS were correlated with the GSR and ASR scores using Spearman correlations. We used the pwr.r.test() function from the pwr package [53] to determine the statistical power of the correlations. Multiple testing corrections for the 16 PGS were performed with the Benjamini and Yekutieli method [25]. We tested for PGS-by-GSR interactions with three linear models and then performed stratified correlations.

The first model included the covariates (age, sex, and autism) as both main effects and interactions with the PGS and GSR scores, as recommended by [26]. The model was specified as: *ASR ∼ PGS* + *GSR* + *PGS × GSR*+*age*+*sex*+*autism*+*PGS ×age*+*PGS ×sex*+*PGS ×autism*+*GSR×age*+*GSR×sex*+*GSR×autism*. For the second model, we wanted to be consistent with the previous analyses. We tested for interactions with the variables adjusted for covariates prior to model input. This model was specified as: *ASR ∼ PGS* + *GSR* + *PGS × ASR*, with the ASR, PGS, and GSR adjusted for age, sex, and autism. For the third model, we binarized the PGS into the upper quartile (coded as 1) and the lower quartile groups (coded as 0), with *N* = 160 in each group and the middle 50% removed. This model was specified as: *ASR ∼ PGSgroup* + *GSR* + *PGSgroup × GSR*, with the ASR and GSR scores adjusted for covariates prior to model input. To further investigate the interactions from the third model, we ran ASR-GSR correlations stratified by PGS groups.

## Data Availability

The SPARK genetic data can be obtained at SFARI Base. The SPARK Research Match data will be available to qualified, approved researchers through SFARI Base upon publication of this article.

https://base.sfari.org

## ACKNOWLEDGEMENTS

We are grateful to our community advisory council, including members Elizabeth Graham, Sascha Klomp, and Jillian Nelson for their feedback throughout the research and writing process. We are also grateful to all the participants and families in SPARK, the SPARK clinical sites, and the SPARK staff. We appreciate obtaining access to genetic and phenotypic data for SPARK data on SFARI Base.

## DATA AND CODE AVAILABILITY

SPARK genetic data can be obtained at SFARI Base: https://base.sfari.org

SPARK Research Match data will be available to qualified approved researchers through SFARI Base after publication of this article. The code for all analyses can be found at https://research-git.uiowa.edu/ michaelson-lab-public/gsr-polygenic-scores

## AUTHOR CONTRIBUTIONS

The study was designed by TRT, JFS, and JJM. The GSR scores were generated by JSY and JFS. The polygenic scores were generated by TRT and JJM. The analyses were performed by TRT, AJT, and JJM. The manuscript writing was done by all authors.

## SUPPLEMENTARY

**Table S1.** Polygenic score associations with the Gender Self Report and Adult Self Report scores using linear regression with covariates. Linear regressions to test for the polygenic (PGS) associations with the Gender Self Report (GSR) and Adult Self Report (ASR) scores with age, sex, and autism as covariates in the model: *phenotype ∼ PGS* +*age*+*sex*+*autism*. The ASR and GSR phenotypes and PGS were not adjusted for covariates prior to model input, but they were Z-scaled (*µ* = 0, *σ* = 1). The regression were ran in the European subset (*N* = 639).PGS abbreviations: **risky** = risky behavior, **NEB** = number of children ever born, **non-het** = non-heterosexual sexual behavior, **SCZ** = schizophrenia.

| phenotype | PGS | PGS $\beta$ (95% CI) | $\beta$ pval | $\beta$ pval FDR |
| --- | --- | --- | --- | --- |
| **ASR Externalizing** | **ADHD** | **0.1 [0.03, 0.17]** | **0.01** | **0.45** |
| **ASR Externalizing** | **anorexia** | **-0.07 [-0.15, 0]** | **0.04** | **>.999** |
| ASR Externalizing | autism | 0.01 [-0.06, 0.08] | 0.80 | >.999 |
| ASR Externalizing | bipolar | 0.01 [-0.06, 0.08] | 0.79 | >.999 |
| ASR Externalizing | cog perf | 0 [-0.08, 0.07] | 0.92 | >.999 |
| ASR Externalizing | depression | 0.05 [-0.03, 0.12] | 0.21 | >.999 |
| ASR Externalizing | edu attain | -0.04 [-0.12, 0.03] | 0.27 | >.999 |
| ASR Externalizing | extraversion | -0.01 [-0.08, 0.06] | 0.82 | >.999 |
| ASR Externalizing | NEB men | 0.02 [-0.05, 0.1] | 0.54 | >.999 |
| ASR Externalizing | NEB women | 0.01 [-0.06, 0.09] | 0.74 | >.999 |
| ASR Externalizing | neuroticism | -0.01 [-0.08, 0.07] | 0.89 | >.999 |
| ASR Externalizing | non-het | 0.01 [-0.06, 0.09] | 0.72 | >.999 |
| ASR Externalizing | OCD | -0.04 [-0.11, 0.03] | 0.30 | >.999 |
| ASR Externalizing | openness | -0.05 [-0.12, 0.02] | 0.17 | >.999 |
| ASR Externalizing | risky | 0.03 [-0.04, 0.11] | 0.37 | >.999 |
| ASR Externalizing | SCZ | -0.06 [-0.13, 0.01] | 0.11 | >.999 |
| ASR Internalizing | ADHD | 0.01 [-0.06, 0.08] | 0.73 | >.999 |
| ASR Internalizing | anorexia | 0 [-0.07, 0.07] | 0.90 | >.999 |
| ASR Internalizing | autism | 0.01 [-0.06, 0.08] | 0.72 | >.999 |
| ASR Internalizing | bipolar | 0.02 [-0.05, 0.09] | 0.58 | >.999 |
| ASR Internalizing | cog perf | 0.02 [-0.05, 0.09] | 0.53 | >.999 |
| **ASR Internalizing** | **depression** | **0.07 [0, 0.14]** | **0.04** | **>.999** |
| ASR Internalizing | edu attain | -0.03 [-0.1, 0.04] | 0.41 | >.999 |
| ASR Internalizing | extraversion | -0.04 [-0.11, 0.03] | 0.29 | >.999 |
| ASR Internalizing | NEB men | -0.03 [-0.1, 0.05] | 0.48 | >.999 |
| ASR Internalizing | NEB women | 0 [-0.07, 0.07] | 0.92 | >.999 |
| **ASR Internalizing** | **neuroticism** | **0.1 [0.03, 0.17]** | **0.01** | **0.32** |
| ASR Internalizing | non-het | 0.02 [-0.05, 0.09] | 0.53 | >.999 |
| ASR Internalizing | OCD | -0.03 [-0.1, 0.04] | 0.37 | >.999 |
| ASR Internalizing | openness | -0.01 [-0.08, 0.06] | 0.81 | >.999 |
| ASR Internalizing | risky | -0.06 [-0.13, 0.01] | 0.08 | >.999 |
| ASR Internalizing | SCZ | -0.01 [-0.08, 0.06] | 0.87 | >.999 |
| GSR Binary | ADHD | 0.02 [-0.04, 0.09] | 0.46 | >.999 |
| GSR Binary | anorexia | -0.01 [-0.07, 0.06] | 0.87 | >.999 |
| GSR Binary | autism | 0.03 [-0.04, 0.09] | 0.45 | >.999 |
| GSR Binary | bipolar | -0.01 [-0.07, 0.06] | 0.84 | >.999 |
| GSR Binary | cog perf | 0.07 [0, 0.13] | 0.06 | >.999 |
| GSR Binary | depression | -0.02 [-0.09, 0.04] | 0.51 | >.999 |
| GSR Binary | edu attain | 0 [-0.06, 0.07] | 0.89 | >.999 |
| GSR Binary | extraversion | 0.01 [-0.06, 0.07] | 0.84 | >.999 |
| GSR Binary | NEB men | -0.01 [-0.07, 0.06] | 0.87 | >.999 |
| GSR Binary | NEB women | 0.04 [-0.03, 0.11] | 0.23 | >.999 |
| GSR Binary | neuroticism | 0.02 [-0.05, 0.08] | 0.65 | >.999 |
| **GSR Binary** | **non-het** | **0.07 [0.01, 0.14]** | **0.03** | **>.999** |
| GSR Binary | OCD | 0.02 [-0.04, 0.09] | 0.51 | >.999 |
| GSR Binary | openness | -0.02 [-0.08, 0.05] | 0.58 | >.999 |
| GSR Binary | risky | -0.02 [-0.08, 0.05] | 0.60 | >.999 |
| GSR Binary | SCZ | 0.03 [-0.04, 0.09] | 0.42 | >.999 |
| GSR Nonbinary | ADHD | -0.02 [-0.08, 0.05] | 0.58 | >.999 |
| GSR Nonbinary | anorexia | 0.03 [-0.03, 0.1] | 0.33 | >.999 |
| GSR Nonbinary | autism | 0.01 [-0.06, 0.07] | 0.80 | >.999 |
| GSR Nonbinary | bipolar | -0.03 [-0.1, 0.03] | 0.33 | >.999 |
| **GSR Nonbinary** | **cog perf** | **0.11 [0.05, 0.18]** | **<.001** | **0.05** |
| GSR Nonbinary | depression | -0.03 [-0.09, 0.04] | 0.38 | >.999 |
| GSR Nonbinary | edu attain | 0.07 [0, 0.13] | 0.05 | >.999 |
| GSR Nonbinary | extraversion | -0.04 [-0.11, 0.02] | 0.20 | >.999 |
| GSR Nonbinary | NEB men | -0.04 [-0.11, 0.02] | 0.19 | >.999 |
| GSR Nonbinary | NEB women | -0.02 [-0.09, 0.04] | 0.54 | >.999 |
| GSR Nonbinary | neuroticism | -0.01 [-0.08, 0.05] | 0.71 | >.999 |
| GSR Nonbinary | non-het | 0.04 [-0.03, 0.11] | 0.22 | >.999 |
| GSR Nonbinary | OCD | -0.01 [-0.07, 0.06] | 0.87 | >.999 |
| GSR Nonbinary | openness | 0.01 [-0.05, 0.08] | 0.66 | >.999 |
| GSR Nonbinary | risky | -0.01 [-0.08, 0.05] | 0.70 | >.999 |
| GSR Nonbinary | SCZ | -0.03 [-0.09, 0.04] | 0.45 | >.999 |

**Figure S1.**
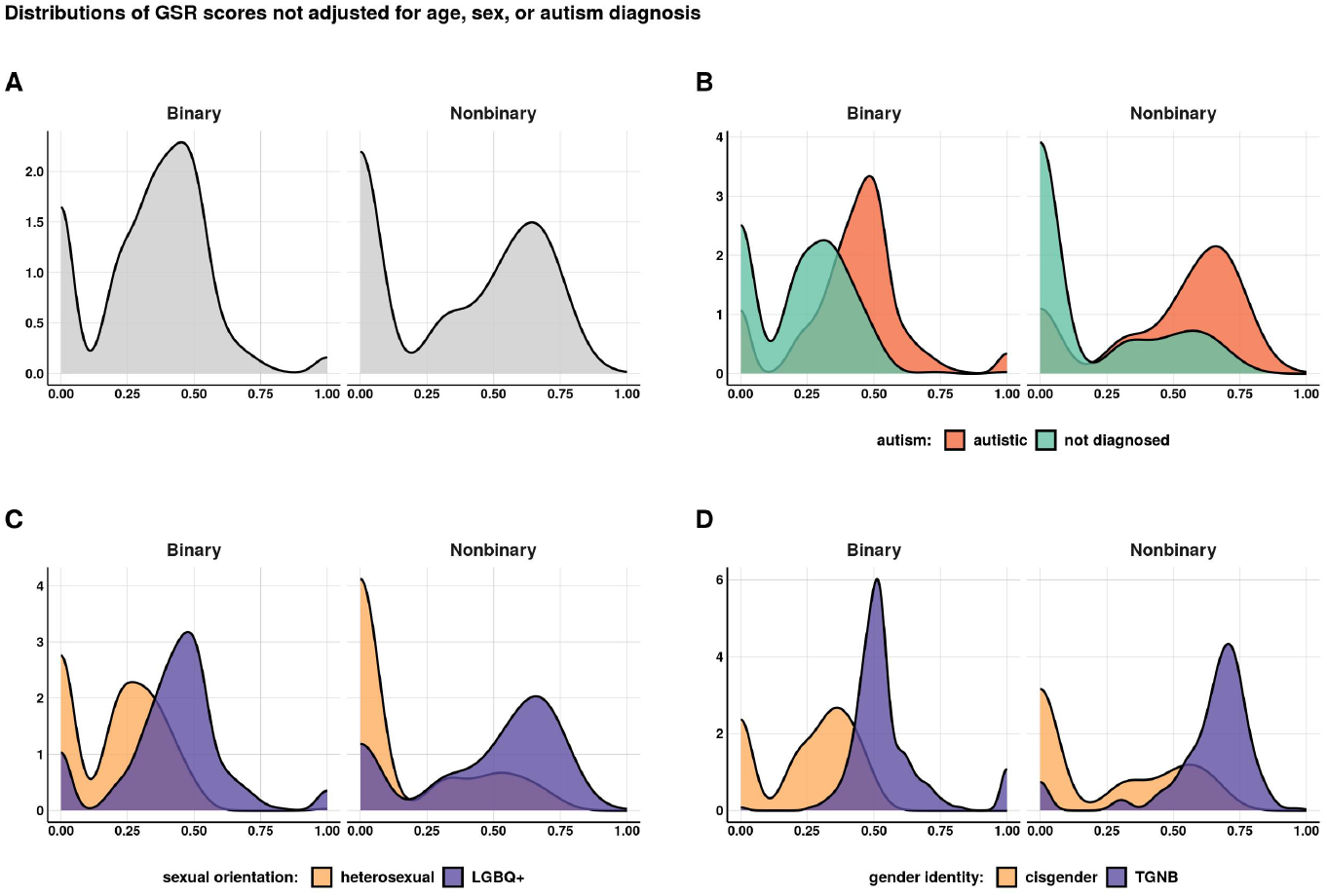
Distributions of the Gender Self Report scores before adjusting for covariates. The Gender Self Report (GSR) scores here are the unadjusted scores from the GSR factor analysis [24].

**Figure S2.**
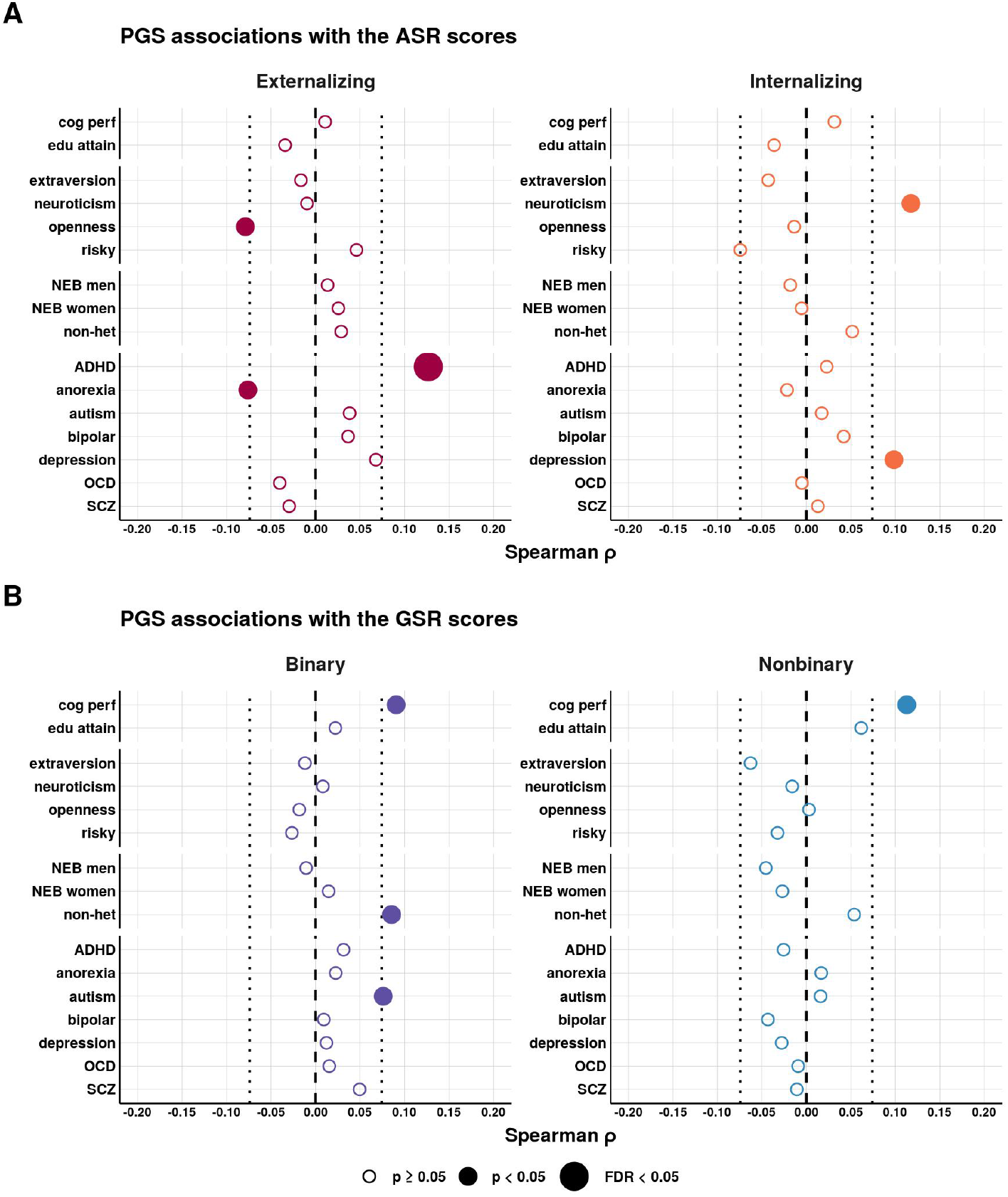
Polygenic score associations with the Gender Self Report and Adult Self Report scores, not filtered by genetic ancestry. Polygenic score (PGS) associations with the **(A)** Adult Self Report (ASR) scores and **(B)** Gender Self Report (GSR) scores. The PGS, as well as the GSR and ASR phenotypes, were adjusted for age, sex, and autism. The correlations shown here were run in the full cohort (*N* = 696). PGS abbreviations: **risky** = risky behavior, **NEB** = number of children ever born, **non-het** = non-heterosexual sexual behavior, **SCZ** = schizophrenia.

**Figure S3.**
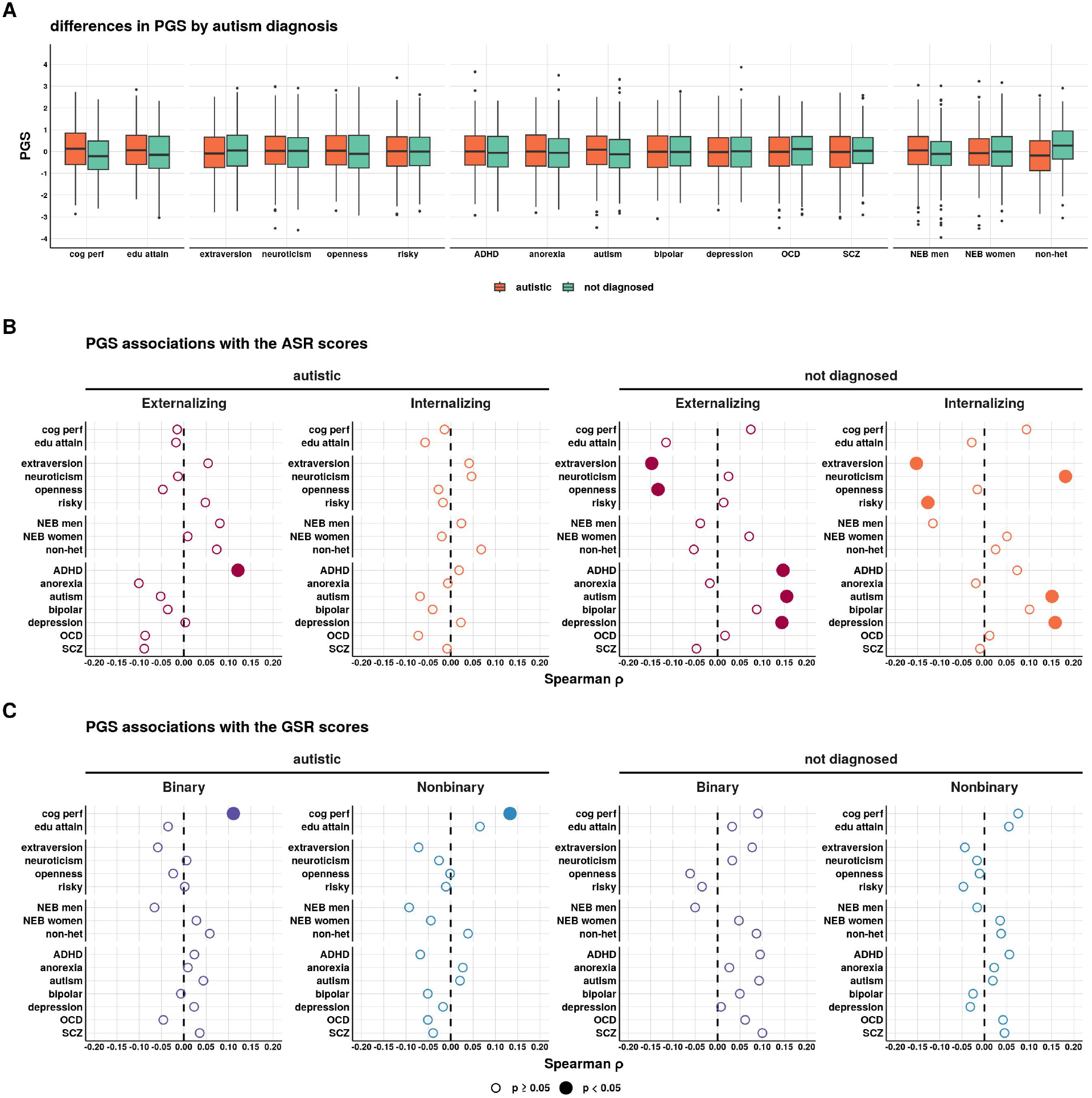
Polygenic score associations with the Gender Self Report and Adult Self Report scores, stratified by autism diagnosis. (A) Polygenic score (PGS) differences by autism diagnosis within those of the European genetic ancestry cluster (Autistic *N* = 376, not diagnosed *N* = 263). PGS correlations with the **(B)** Adult Self Report (ASR) scores and **(C)** Gender Self Report (GSR) scores. The PGS, as well as the GSR and ASR phenotypes, were not adjusted for age or sex. Multiple testing corrections for the 16 PGS were performed with the Benjamini and Yekutieli method [25]. PGS abbreviations: **risky** = risky behavior, **NEB** = number of children ever born, **non-het** = non-heterosexual sexual behavior, **SCZ**= schizophrenia.

**Figure S4.**
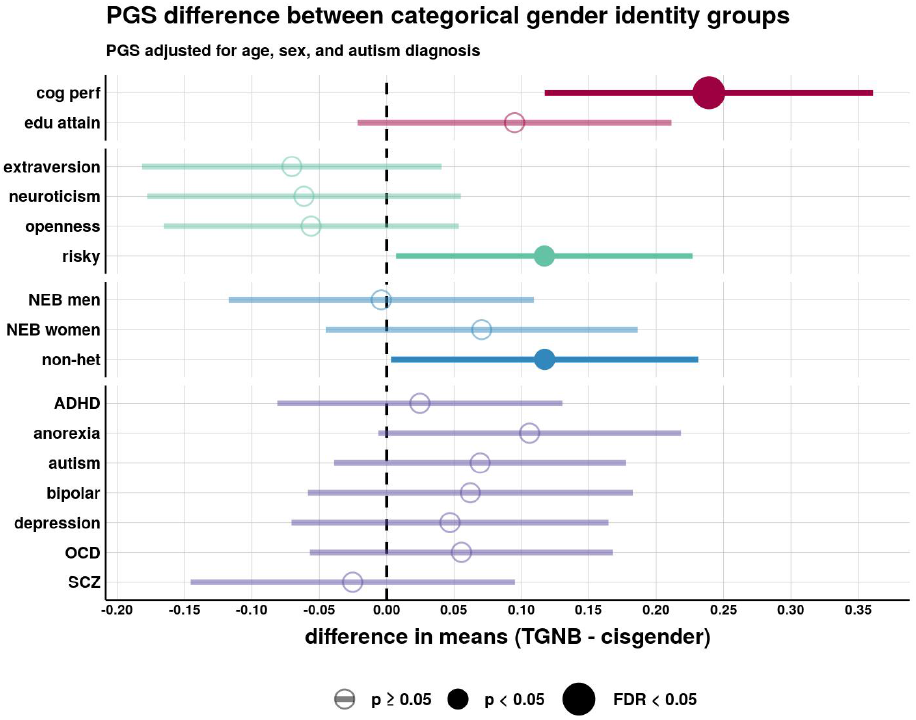
Replication of polygenic score associations with categorical gender identity in a larger sample, not filtered on genetic ancestry. Polygenic score (PGS) difference in means (t-tests) between gender identity groups in the larger cohort (*N* = 5, 388) with all samples. The two gender identity groups are cisgender (*N* = 5, 089) versus transgender and gender nonbinary (TGNB, *N* = 299). The PGS were adjusted for age, sex, and autism prior to performing the t-tests. Multiple testing corrections for the 16 PGS were performed with the Benjamini and Yekutieli method [25]). PGS abbreviations: **risky** = risky behavior, **NEB** = number of children ever born, **non-het** = non-heterosexual sexual behavior, **SCZ** = schizophrenia.

**Figure S5.**
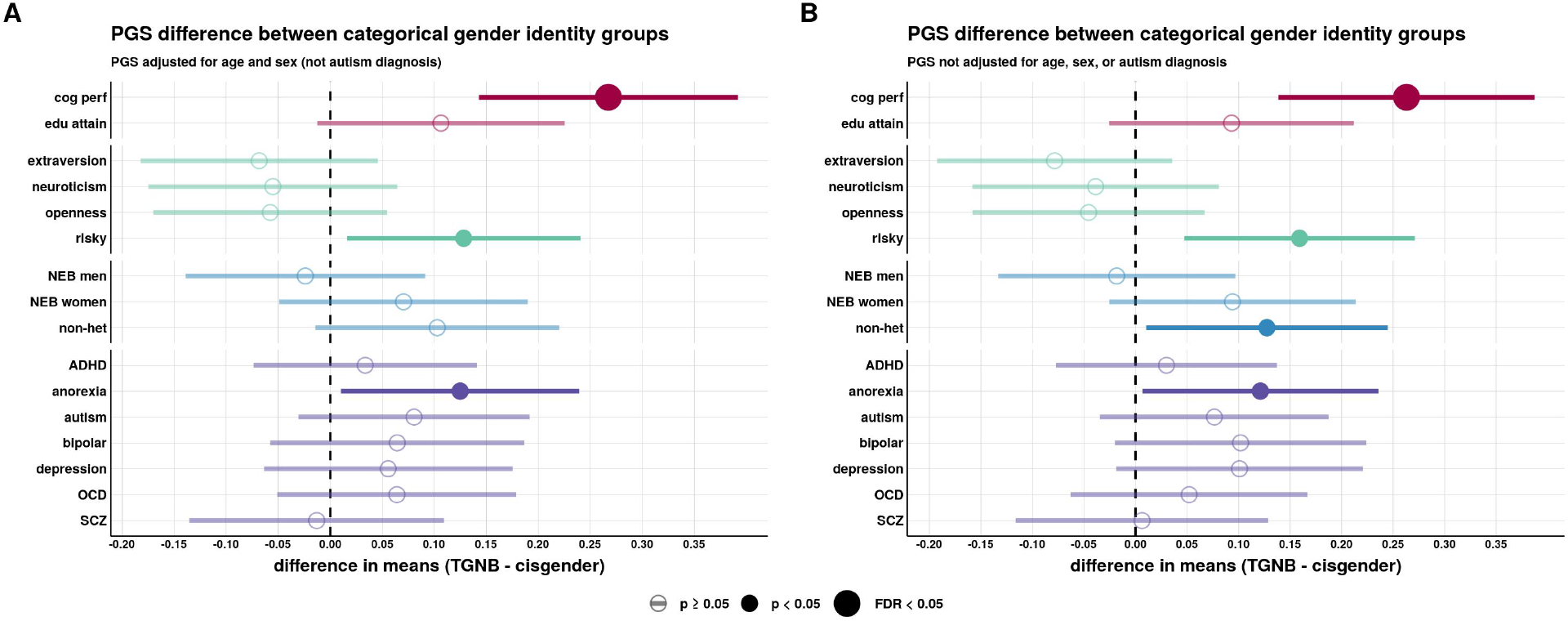
Replication of polygenic score associations with categorical gender identity in a larger sample with the polygenic scores not adjusted for age, sex, or autism. Polygenic score (PGS) difference in means (t-tests) between gender identity groups in the larger cohort (*N* = 5, 165) of European genetic ancestry. The two gender identity groups are cisgender (*N* = 4, 879) versus transgender and gender nonbinary (TGNB, *N* = 286). **(A)** shows the results for the PGS adjusted for age and sex (not autism). **(B)** shows the results with the PGS not adjusted for age, sex, nor autism. Multiple testing corrections for the 16 PGS were performed with the Benjamini and Yekutieli method [25]. PGS abbreviations: **risky** = risky behavior, **NEB** = number of children ever born, **non-het** = non-heterosexual sexual behavior, **SCZ** = schizophrenia.

**Figure S6.**
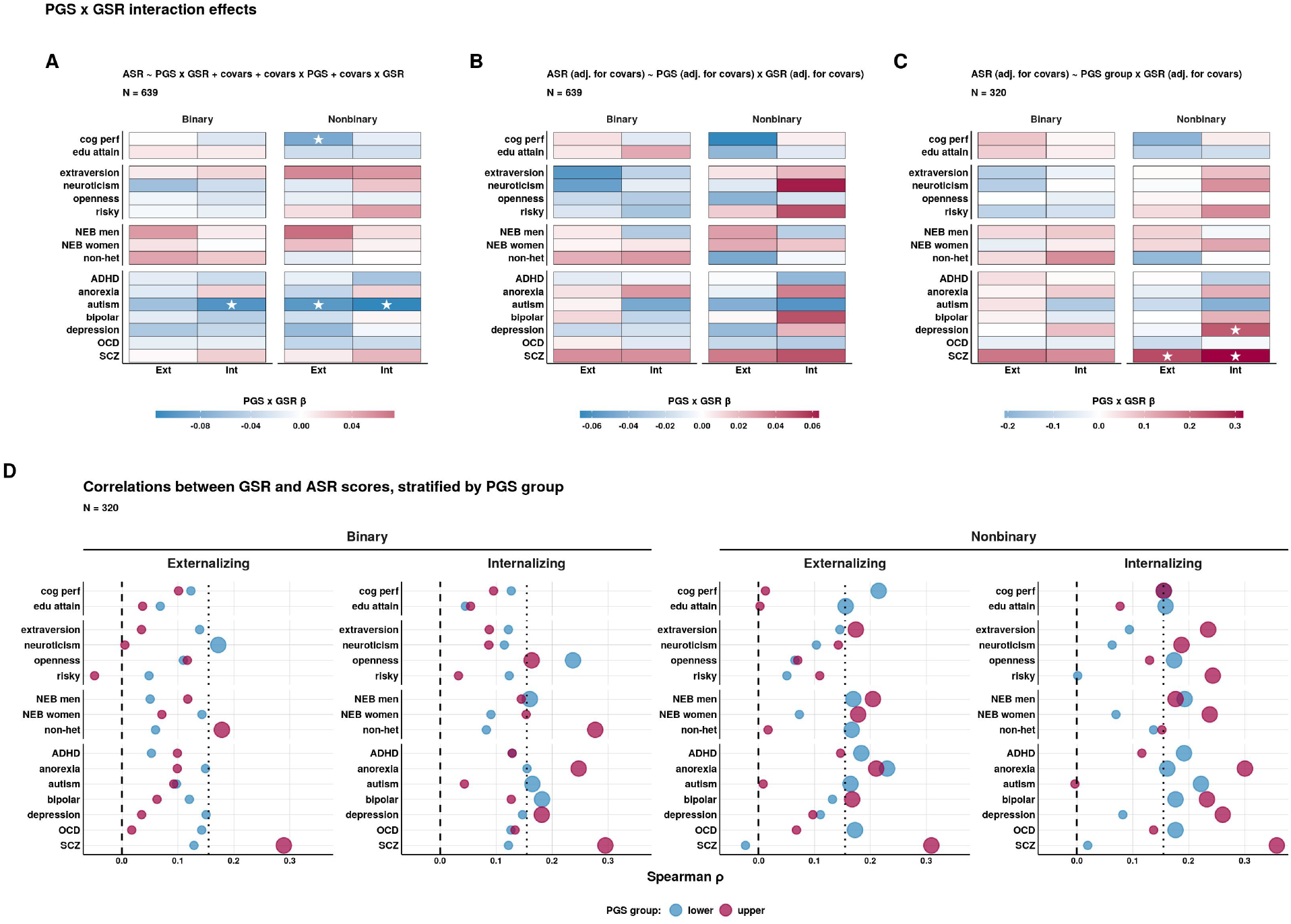
Interaction effects between the Gender Self Report and polygenic scores on the Adult Self Report. Interactions between the Gender Self Report (GSR) scores and the polygenic scores (PGS) on the Adult Self Report (ASR) scores were tested with three models. All variables were Z-scaled prior to model input. The fill color is the *β* estimate for the PGS-by-GSR interaction term from the linear model. Nominally significant interaction terms are indicated with one star. No interactions are significant after multiple testing corrections with the Benjamini and Yekutieli method [25]. **(A)** As recommended by [26], this model included the covariates (age, sex, and autism) as both main effects and interactions with the PGS and GSR scores. *ASR ∼ PGS* + *GSR* + *PGS × GSR* + *age* + *sex* + *autism* + *PGS × age* + *PGS × sex* + *PGS × autism* + *GSR × age* + *GSR × sex* + *GSR × autism*. **(B)** To be consistent with the previous analysis, we tested for interactions with the variables adjusted for covariates prior to model input. Specifically, the PGS, GSR, and ASR scores were adjusted for age, sex, and autism. The interaction model was thus *ASR ∼ PGS* + *GSR* + *PGS × ASR*. **(C)** We binarized the PGS: the upper 75th quartile (coded as 1) and the lower 25th quartile groups (coded as 0), with *N* = 160 in each group and the middle 50% removed. The interaction model was thus *ASR ∼ PGSgroup* + *GSR* + *PGSgroup × GSR*, with the ASR and GSR scores adjusted for covariates prior to model input. **(D)** Spearman correlations of the GSR scores with the ASR scores stratified by PGS group. The ASR and GSR scores were adjusted for covariates prior to running the correlations. PGS abbreviations: **risky** = risky behavior, **NEB** = number of children ever born, **non-het** = non-heterosexual sexual behavior, **SCZ** = schizophrenia.

## Public summary

The way we act (behavior) is influenced by how our brains grow and function. Some of the ways our brains grow and function are influenced by our genes (DNA). Everyone has slightly different versions of DNA. This is a normal part of human diversity. Some of these DNA differences lead to differences in our brains. Brain differences can lead to behavior differences like in personality, intelligence, or mental health.

In this study, we asked whether our DNA is involved in gender identity and gender expression. We use the term “gender diversity” to mean differences in gender identity or gender expression. People with higher gender diversity are more likely to be transgender and/or nonbinary, although cisgender people can also have differences in gender expression. People with higher gender diversity are also more likely to be autistic, so we conducted this study with the help of SPARK participants (SPARK is the largest study of autism). Approximately half of our study participants were autistic adults, and the others were not autistic but do have an immediate relative who is autistic.

### What we found

We found that thousands of DNA differences, when combined, are linked to differences in gender diversity. Specifically, we found that DNA differences linked to higher intelligence were also linked to higher gender diversity. We need to do more research to understand why DNA differences linked to higher intelligence are also linked to higher gender diversity.

### What our study does not show

We did not identify or attempt to identify a “transgender” or “nonbinary” gene. We cannot predict a person’s gender diversity from their DNA. We found very little evidence that the DNA differences linked to major psychiatric conditions are also strongly linked to gender diversity. Larger studies in the future may be able to identify weaker effects, but our study does not support a strong genetic connection between psychiatric conditions and gender diversity. Gender diversity is not purely genetic, but genetic factors do play a role.

### Why this study is important

This is one of the first genetic studies of gender diversity. Many people say that gender is a purely social construct, with no biological factors involved. Our results show that the DNA differences linked to higher intelligence are also linked to higher gender diversity. Ultimately, we believe this line of research will advance the health of gender diverse people through a higher understanding of how genetics interact with gender diversity in determining health outcomes.

